# Effect of Virtually Led Value-Based Preoperative Assessment on Safety, Efficiency, and Patient and Professional Satisfaction

**DOI:** 10.1101/2025.03.11.25323740

**Authors:** José Luis Gracia Martínez, Miguel Ángel Morales Coca, Marta del Olmo Rodríguez, Pablo Vigoa, Jorge Martínez Gómez, Jorge Short Apellaniz, Catalina Paredes Coronel, Marco Antonio Villegas García, Bernadette Pfang, Juan José Serrano, Javier Arcos, Cristina Caramés Sánchez, Juan Antonio Álvaro de la Parra

## Abstract

**Background:** Increasing demand for elective surgery makes optimizing preoperative assessment a priority. Value-based healthcare aims to provide the highest value for patients at the lowest possible cost through various mechanisms including reorganizing care into integrated practice units (IPUs). However, few studies have analyzed the effectiveness of implementing virtually led IPUs for preoperative assessment.

**Methods:** We performed a retrospective observational cohort study including patients undergoing elective surgery at a teaching hospital in Madrid, Spain from January 1^st^, 2018, to December 31^st^, 2023, analyzing changes in surgical complications, efficiency, and patient satisfaction between the preimplementation (2018-2019) and postimplementation (2020-2023) periods. Anesthesiologist satisfaction with virtual assessment was described. During the postimplementation period, preoperative assessment was reorganized as a virtually led IPU. At the IPU appointment, preoperative testing and physical (including airway) examination was performed by a nurse anesthesiologist. Results were uploaded to the electronic health record and asynchronous virtual anesthesiologist assessment using a store-and-forward approach was performed. Digital patient education was carried out over the Patient Portal mobile application.

**Results:** A total of 40,233 surgical procedures were included, of which 31,259 were from the postintervention period. During the postintervention period, no increase in surgical complications was observed, while same-day cancellations decreased from 4.3% to 2.8% of total procedures (P<0.001). Overall process time did not increase, despite the rising number of surgical procedures per year. Patient satisfaction improved. Median time to complete anesthesiologist assessment was significantly lower for virtual assessment (4.5 versus 10 minutes (P<0.001), signifying estimated time savings of 716 person-hours per year. Anesthesiologists agreed that virtual assessment was more efficient that in-person evaluation, and half of participants agreed that virtual preoperative care improved work-life balance and reduced burn-out.

**Conclusions:** A digitally enhanced value-based model of preoperative care can improve efficiency and satisfaction metrics, reducing unnecessary costs and potentially improving quality of care.

## INTRODUCTION

Preoperative assessment is a key component for ensuring safe, sustainable surgical care^1^. Preanesthesia evaluation clinics (PECs) for outpatient anesthetist-led preoperative assessment have demonstrated reductions in same-day surgical cancellations, surgical delays, and healthcare-related expenditure^2–4^. However, while PECs are increasingly common, the best model of PEC organization is unclear. The increasing demand for elective surgery in developed countries makes optimization of preoperative assessment an international priority for anesthesiologists, surgeons, and healthcare managers^5,6^.

Value-based healthcare emerged at the beginning of the century as a strategy to solve current problems in healthcare by focusing on delivering the highest value for patients at the lowest possible cost^7^. One of the main components of value-based care is the integration of care delivery by reorganizing care into integrated practice units (IPUs), in which different health professionals and services work together in the same physical space to provide patient-centered care and sharing responsibility for costs and outcomes^8,9^. While in other specialties, IPUs have demonstrated improved patient satisfaction, reduced waiting times for patients, and better outcomes^1011–13^, scarce literature exists on the implementation of integrated care delivery in preoperative care assessment.

On the other hand, the increasing importance of information and communication technology in medicine has led to a growing use of preoperative telemedicine^14^. Digital interventions have proven a useful way of improving patient education prior to surgery and providing patient-centered care^15^. Available evidence indicates that virtual preoperative assessment is comparable to in-person evaluation regarding patient safety, patient experience, and efficiency^14,16,17^. However, the effectiveness of combining virtual care and IPUs in preoperative assessment has not been described.

This study evaluates the effects of redesigning an outpatient preanesthesia evaluation clinic in Madrid, Spain, as a virtually led IPU. Using six years of data, we described changes in patient safety, patient satisfaction, and same-day surgical cancellations before and after implementation. We also performed a subanalysis of efficiency metrics and healthcare professional experience comparing virtual anesthesiology assessment with traditional, in-person evaluation.

## METHODS

### Study design

A retrospective before-after observational study was carried out including pseudoanonymized data from all elective surgical procedures performed at the General Villalba Hospital (Madrid, Spain) during a six-year period. The pre-implementation period was defined as January 1^st^, 2018, to December 31^st^, 2019. The post-implementation period was defined from January 1^st^, 2020, to December 31^st^, 2023. We excluded urgent and emergent procedures, as well as procedures taking place in patients younger than 18 years.

### Intervention

The intervention was designed from September – December 2019 to address the study hospital’s increasing clinical burden of preoperative assessment. An anesthetist-led multidisciplinary team was formed to identify areas for improvement, including anesthesiologists, nurse anesthesiologists, members of the information technology and systems department, and hospital managers. Over a series of brainstorming sessions, “logistical pitfalls” along the preoperative patient journey were identified (Figure 1). To remediate these problems, a new patient workflow was designed, including a digital patient-administered questionnaire before the PEC appointment, IPU with nurse practitioner-led preanesthesia evaluation, and virtual anesthetist supervision for most patients (Figure 2). Before consultation, patients fill out a preanesthesia questionnaire using a webapp (the Quirónsalud Patient Portal), answering standardized questions about preexisting conditions and current medications.

**Figure 1:**
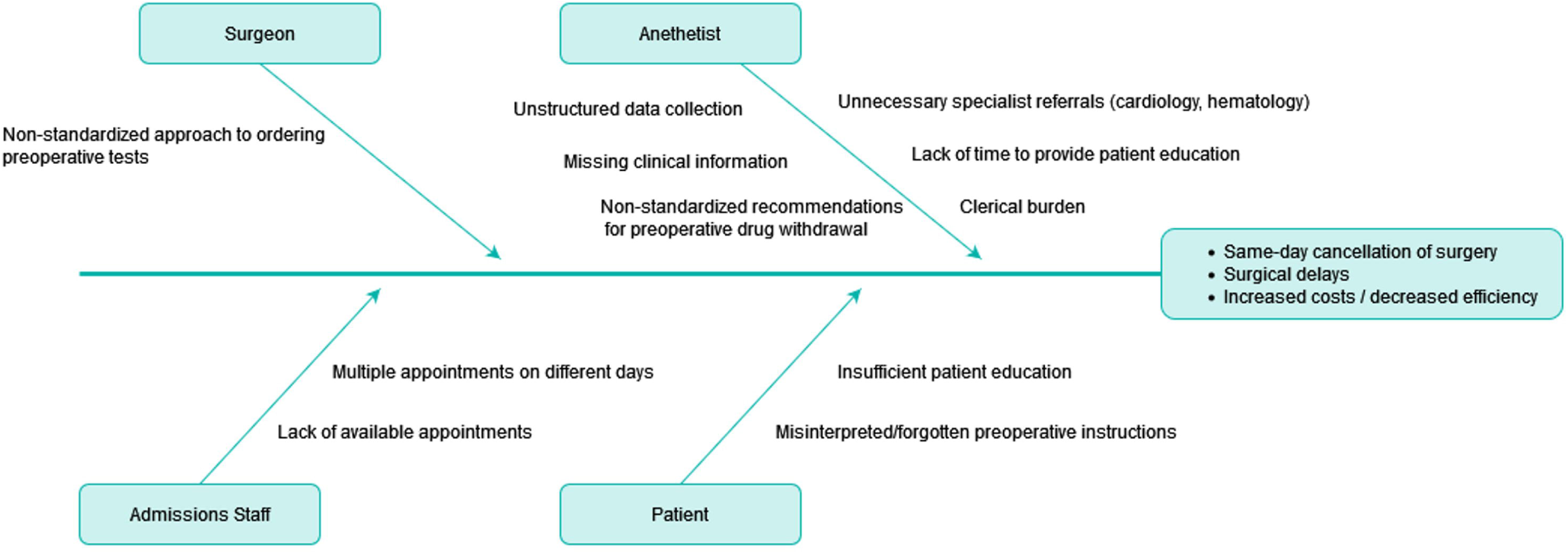
Cause and effect diagram highlighting causes of inefficiency in preoperative assessment workflow. We identified principal causes behind inefficiencies of the traditional PEC model, including non-standardization, unstructured data collection, non-standardized preoperative recommendations, and lack of patient education, leading to preventable same-day cancellations of surgery, surgical delays, and increased costs.

**Figure 2:**
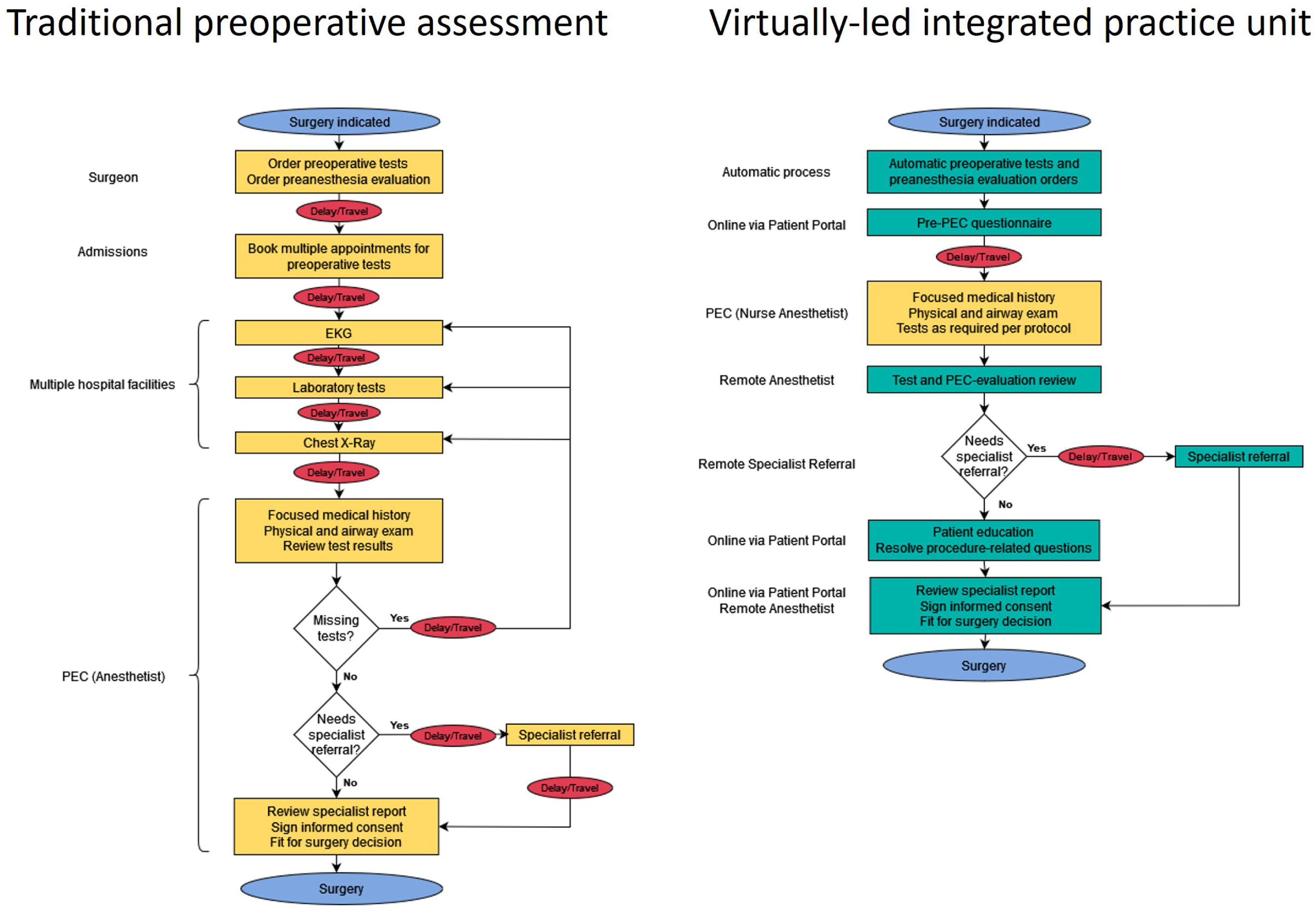
Process map comparing traditional preoperative evaluation clinic workflow and virtually-led integrated practice unit (IPU) preoperative assessment. Process map of traditional preoperative assessment compared to a virtually-led integrated practice unit for preoperative assessment

A previously described algorithm was developed and integrated within the EHR to automatically place preoperative test orders based on surgical and patient characteristics^18^. The aim of the algorithm was to reduce unnecessary preoperative testing in line with patient safety initiatives such as “Choosing Wisely”^19,20^. We created an integrated practice unit (IPU) to unify testing and nurse-led preanesthesia evaluation on the same day, in the same space, reducing patient travel and wait times. Nurse anesthetists receive specific training on focused clinical history and physical exam, using a checklist integrated with the EHR to ensure all necessary items are covered. We developed a remote airway exam protocol, permitting virtual airway exam through a series of predetermined photographs uploaded to the EHR. At the end of the IPU encounter, all patients are discharged for virtual asynchronous (store-and-forward) anesthetist review. For patients discharged for virtual review, anesthetists review the nurse reports and test results over the EHR, and either digitally approve patients for surgery or – exceptionally – order additional tests, specialist referrals, or an in-person anesthetist evaluation. Further patient education is provided through the Patient Portal, including educational videos on the benefits and potential risks of the type of anesthesia that will be used during their procedure. Patients are able to sign informed consent electronically over the Patient Portal after completing the educational material.

### Objectives and measurements

#### Primary objectives

The primary objective of this study was to describe and analyze changes in surgical complications, efficiency metrics, and patient satisfaction after reorganizing the study hospital’s outpatient PEC as a virtually led IPU.

#### Safety metrics

Surgical complications were used to estimate the effect of the initiative on patient safety. We analyzed Agency for Healthcare Research and Quality Patient Safety Indicators (AHRQ-PSI) recorded as part of annual internal audit data (rates of postoperative bleeding, respiratory failure, pulmonary thromboembolism, deep vein thrombosis, sepsis, and wound dehiscence) for the pre- and post-implementation periods.

#### Efficiency measures

Efficiency measures included in this study included rates of same-day cancellation and overall process times. To calculate rates of same-day surgical cancellation, we excluded 2020 to avoid pandemic-related bias. Same-day cancellation data was taken from the hospital’s surgical database.

To calculate process times, we randomly selected a sample of 381 patients from both groups, stratified by age and sex. Overall process time was defined as the time from the referral for preoperative evaluation to the date in which patients underwent surgery.

#### Patient satisfaction

To describe patient satisfaction, we analyzed differences in the NPS (Net Promoter Score)^21^. NPS surveys were randomly administered to patients attending the study hospital PEC during the pre- and post-intervention periods. Scores ranged from -100 to 100.

### Secondary objectives

Secondary objectives were to describe demographic differences in the postimplementation period between patients undergoing virtually led assessment and those undergoing additional in-person anesthetist assessment. We also analyzed the difference in the time that anesthesiologists took to complete virtually led and in-person assessment, and the differences in process time between the IPU encounter and anesthesiologist assessment when this was performed virtually and in-person, respectively.

To evaluate anesthesiologist satisfaction with virtually led assessment, after performing a qualitative literature review to identify important aspects of clinician experience in virtual preoperative evaluation, we designed a 12-item survey which was sent via email to anesthesiologists at the study hospital. Responses were recorded anonymously. The full survey is available as Appendix 1.

### Statistical analysis

Statistical analysis was performed using Python v 3.10. The Kolmogorov-Smirnov test was performed to determine the distribution of the different variables. Continuous variables were expressed as mean (SD) or median (IQR), according to distribution. Categorical variables were expressed as number (percentage). The Mann-Whitney U test was used to compare differences in non-normal continuous variables between the study and control groups. The Chi Squared test was used to compare differences in categorical variables. A P-value of <0.05 was deemed statistically significant.

### Ethics and reporting standards

The study was carried out according to the principles set forth in the Declaration of Helsinki. All data were pseudonymized. The study design was approved by the institutional ethics committee (ethics approval code: EO084-24). Due to the retrospective nature of the study, the requirement for informed consent was waived. As a quality improvement initiative, SQUIRE 2.0 guidelines ^22^ were followed when reporting the study.

## RESULTS

### Comparison of preintervention and postintervention outcomes

During the preintervention period, a total of 8,974 patients underwent preoperative assessment at the participating center, compared to 31,259 patients in the postintervention period. Median age was 58 years (27-84), and 46.5% of patients were female. Differences in demographic characteristics of patients between periods are summarized in Table 1.

**Table 1.** Differences in demographic characteristics, adverse preoperative events, patient satisfaction and same-day cancellations between the preintervention and postintervention periods.

|  | Preintervention period<br>(2018-2019)<br>N=8,974 | Postintervention<br>period (2020-2021)<br>N=31,259 | P-Value |
| --- | --- | --- | --- |
| Age, years (median (IQR)) | 57 (25 – 85) | 59 (27 – 84) | 0.003 |
| Female sex (n, %) | 4,316 (48.1%) | 14,382 (46.0%) | 0.007 |
| Postoperative bleeding* | 18 (0.4%) | 36 (0.3%) | 0.83 |
| Postoperative respiratory failure* | 10 (0.3%) | 19 (0.3%) | 0.62 |
| Pulmonary thromboembolism/Deep vein thrombosis* | 8 (0.2%) | 15 (0.1%) | 0.69 |
| Postoperative sepsis* | 1 (0.1%) | 8 (0.1%) | 0.56 |
| Wound dehiscence* | 4 (0.4%) | 10 (0.4%) | 0.92 |
| Overall NPS | 69.7 | 73.4 | N/A |
| Same-day cancellations | 384 (4.3%) | 775 (2.8%)** | <0.001 |

\*Agency for Healthcare Research and Quality Patient Safety Indicators (AHRQ-PSI) recorded as part of annual internal audit data (rates of postoperative bleeding, respiratory failure, pulmonary thromboembolism, deep vein thrombosis, sepsis, and wound dehiscence). For complete data including sample size and inclusion criteria, see Supplement 2.
\*\* Data from 2020 was excluded from analysis.

Regarding patient safety, no differences in adverse perioperative events were observed between both periods (Table 1). Patient satisfaction increased for the postintervention period, from a NPS of 69.7 to 73.4. Same-day surgical cancelations decreased significantly from 4.3% to 2.8% of overall procedures in the postintervention period (p <0.001). No differences were observed in overall process time despite the increase in total surgical procedures per year.

### Comparison of virtual and in-person anesthesia assessment

We performed a comparison of virtual and in-person anesthesia evaluation during the postintervention period. Median age of patients in the virtual assessment group was almost 10 years younger than that of patients in the in-person group (53 years (21 - 81) versus 62 (20 – 84), P<0.001). Time from the IPU visit to completed anesthesia evaluation was significantly shorter for patients in the virtual assessment group (4 (2-19) days versus 9 (2-25) days, P <0.001).

Regarding anesthesiologist perspectives on the virtually led model of care, 8 (80%) of 10 eligible consultant anesthesiologists answered the survey. Full results are presented in Table 2. Virtual evaluation was estimated to take a median of 4.5 minutes (range 2.5 - 10 minutes), while in-person assessment was estimated to take a median of 10 minutes (range 5 - 12 minutes), signifying estimated time savings of 716 person-hours per year. Discrepancies were observed regarding anesthesiologist perception of the quality of care provided by virtual care, while most participants agreed that virtual care improved efficacy. Half of participants agreed or strongly agreed that virtual preoperative evaluation improved work-life balance and reduced burn-out. For example, some participants used down-time during on-call hours to complete virtual assessments.

**Table 2.** Results of survey on anesthesiologist perception of virtual preoperative assessment.

| QUESTION | ANSWER | N ( %) |
| --- | --- | --- |
| **Sociodemographic data** |  |  |
| Sex | Female | 3 (37.5%) |
|  | Male | 5 (62.5%) |
| Years working as consultant in anaesthesiology | 0-2 | 0 (0%) |
|  | 3-5 | 0 (0%) |
|  | 5-9 | 2 (25%) |
|  | >10 | 6 (75%) |
| Years of experience in virtual preoperative assessment | 1 | 0 (0%) |
|  | 2 | 0 (0%) |
|  | 3 | 3 (37.5%) |
|  | >4 | 5 (62.5%) |
| **Quality, efficacy, and professional experience of virtual preoperative assessment** |  |  |
| Virtual assessment presents similar quality to in-person assessment | Completely disagree | 3 (37.5%) |
|  | Somewhat disagree | 1 (12.5%) |
|  | Neither agree nor disagree | 0 (0%) |
|  | Somewhat agree | 3 (37.5%) |
|  | Completely agree | 1 (12.5%) |
| Virtual assessment is associated with high patient satisfaction | Completely disagree | 0 (0%) |
|  | Somewhat disagree | 3 (37.5%) |
|  | Neither agree nor disagree | 0 (0%) |
|  | Somewhat agree | 3 (37.5%) |
|  | Completely agree | 2 (25%) |
| Virtual assessment reduces the quality of the doctor-patient relationship | Completely disagree | 2 (25%) |
|  | Somewhat disagree | 1 (12.5%) |
|  | Neither agree nor disagree | 2 (25%) |
|  | Somewhat agree | 1 (12.5%) |
|  | Completely agree | 2 (25%) |
| Virtual assessment improves process efficiency | Completely disagree | 1 (12.5%) |
|  | Somewhat disagree | 0 (0%) |
|  | Neither agree nor disagree | 1 (12.5%) |
|  | Somewhat agree | 2 (25%) |
|  | Completely agree | 4 (50%) |
| Virtual assessment improves work-life balance | Completely disagree | 1 (12.5%) |
|  | Somewhat disagree | 1 (12.5%) |
|  | Neither agree nor disagree | 2 (25%) |
|  | Somewhat agree | 0 (0%) |
|  | Completely agree | 4 (50%) |
| Virtual assessment reduces burnout | Completely disagree | 2 (25%) |
|  | Somewhat disagree | 0 (0%) |
|  | Neither agree nor disagree | 1 (12.5%) |
|  | Somewhat agree | 2 (25%) |
|  | Completely agree | 3 (37.5%) |

## DISCUSSION

Our study analyzes six years of data from a teaching hospital in Madrid, Spain, to describe changes in surgical safety metrics, efficiency, and patient satisfaction after implementing a virtually led value-based model of preoperative assessment. No increase in postoperative adverse events or overall process time was observed during the postimplementation period, despite increasing surgical volumes, while same-day cancellations decreased significantly and patient satisfaction scores for preoperative care improved. Upon analyzing postimplementation data, virtually led care was associated with lower process times and patient assessment times. Anesthesiologists generally agreed that virtual preoperative assessment increased efficiency and work-life balance, but opinions were divided as to whether virtual care influenced the quality of care and the clinician-patient relationship.

Many healthcare systems worldwide are investing in value-based care as a potential solution towards delivering high quality, sustainable healthcare^23–26^. The principle behind value-based care is to provide the highest value to patients at the lowest possible cost^7^. In this regard, reorganizing preoperative care as a virtually led integrated practice unit has demonstrated several benefits, for patients and healthcare professionals alike. On the one hand, the new model of care ensures that most patients scheduled for surgery only have to visit the hospital once in order to complete required testing and physical examination, eliminating unnecessary travel and healthcare-related disruptions to daily life. Providing digital patient education over a secure mobile application prior to signing informed consent could theoretically improve patient engagement, which has demonstrated an association with improved clinical outcomes^27^. Likewise, asynchronous virtual anesthesiologist assessment following a “store-and-forward” model decreases the time that anesthesiologists have to dedicate to preoperative assessment by more than 50%, increasing available operating room time and time for other professional activities such as education and research. In this regard, half of surveyed anesthesiologists agreed that virtual assessment improved work-life balance, and – interestingly – reduced professional burn-out.

Since the COVID-19 pandemic, several initiatives featuring virtual preoperative assessment have been reported, demonstrating high overall patient acceptance and similar patient safety results than compared to traditional, in-person care^14,28^. However, to the best of our knowledge, this is the first report of a store-and-forward approach to virtual anesthesiologist assessment. Patients responded favorably to the initiative, as demonstrated by increased Net Promoter Scores during the postimplementation period, while no increase in adverse postoperative events was observed. Most anesthesiologists agreed that patients were satisfied with virtual assessment. However, half of respondents did not agree that the quality of virtual care was equal to that of in-person evaluation. The small sample size of respondents limits the validity of our findings, and future qualitative research is necessary to confirm and further explore these results.

Despite the volume of total surgical procedures per year almost doubling during the postintervention period, no increase in overall process times was observed, which we hypothesize could be due in part to increased efficiency of preoperative assessment after reorganization as an IPU. At the same time, a significant reduction in same-day cancellations was observed, indicating that the new model of care was successful in optimizing patient preparedness for surgery. Cancellation on the day of surgery is an important source of unnecessary healthcare expenditure and contributes to backlogs for elective procedures, with between 10-20% of cancellations due to inadequate preoperative workup^29–32^. Thus, we consider that implementing a virtually led IPU could increase value not only for patients, but also for healthcare systems struggling to improve efficiency of perioperative management and reduce surgical waiting times.

Our study has several limitations. Firstly, its retrospective nature and non-randomized design could potentially lead to biased results. However, data collection during the study period was performed homogeneously and study variables such as patient safety data were calculated on a yearly basis using official definitions by independent departments, reducing the risk of reporting bias. Also, the virtually led IPU was implemented for all patients during the second period of the study, and so changes in variables such as NPS can be attributable, at least in part, to the new model of care. At the same time, although we analyzed anesthesiologist experience, we did not include the perspectives of nurse anesthesiologists operating the IPU, which is a field for further research.

## CONCLUSIONS

Reorganizing preoperative care as a virtually led IPU using a store-and-forward approach is associated with improved patient experience, reduced assessment times, and lower rates of same-day surgical cancellations, without negative effects on patient safety or overall process times despite an increased volume of surgical procedures. A digitally enhanced value-based model of preoperative care can improve efficiency and satisfaction metrics, reducing unnecessary costs and potentially improving quality of care. Further studies are necessary to evaluate anesthesiologist perception of quality of virtual care compared to traditional, in-person preoperative assessment.

## Supporting information

Squire Checklist

## Data Availability

All data produced in the present study are available upon reasonable request to the authors

## Acknowledgements

We would like to thank Juan José Serrano for his help with data acquisition, and the members of the anesthesiology department of the General Villalba Hospital for their enthusiasm supporting this project.

## Summary statement

not applicable

