## Appendix 1 for "Effect of Virtually Led Value-Based Preoperative Assessment on Safety, Efficiency, and Patient and Professional Satisfaction"

Supplementary document 1

Survey for evaluation of anesthesiologist experience with virtual preoperative assessment

1) Male/Female
2) How many years of experience working as a consultant in anesthesiology?
3) How many years of experience with virtual preoperative assessment?
4) On average, how many minutes does it take you
5) to perform one (1) virtual preoperative assessment?
6) On average, how many minutes does it take you to perform one (1) in-person preoperative assessment?
7) The quality of care provided through virtual assessment is equal to that of in-person care.
8) Generally speaking, patients are satisfied with virtual preoperative assessment.
9) Virtual preoperative assessment reduces the quality of the relationship between anesthesiologists and patients.
10) Virtual preoperative assessment improves process efficiency.
11) Virtual preoperative assessment improves my work-life balance.
12) Virtual preoperative assessment helps me to reduce professional burn-out.
13) Other comments (free text).
